# Fecal microbiome predicts treatment response after the initiation of semaglutide or empagliflozin uptake

**DOI:** 10.1101/2024.07.19.24310611

**Authors:** Annabel Klemets, Ingrid Reppo, Kertu Liis Krigul, Vallo Volke, Oliver Aasmets, Elin Org

**Author notes:** These authors contributed equally (Oliver Aasmets, Elin Org).

## Abstract

**Context:** The gut microbiome has been shown to be in a bidirectional interaction with type 2 diabetes medications that have been in clinical use for several decades. However, the bidirectional effects of novel type 2 diabetes drugs semaglutide, empagliflozin, and the gut microbiome have yet to be clearly described.

**Objective:** We investigate the effect of semaglutide and empagliflozin initiation on the gut microbiome of type 2 diabetes patients. In addition, we analyze whether the pre-treatment gut microbiome can predict the treatment efficacy.

**Methods:** Gut microbiome fecal samples donated at four timepoints (Baseline, Month 1, Month 3; Month 12) were studied using 16S ribosomal RNA gene sequencing and analysis. Subjects additionally donated plasma and urine samples for quantitative measurement of clinical markers before treatment initiation and at Months 3 and 12. Repeated measures ANOVA paired with paired t-tests were used to analyze the effects of drug initiation on the gut microbiome. Pearson correlation was used to identify microbial features associated with the change in clinical parameters.

**Results:** Semaglutide and empagliflozin use is associated with changes in the gut microbiome after treatment initiation, but changes in microbial diversity were not detected. The baseline gut microbiome predicted changes in glycohemoglobin for semaglutide and empagliflozin users.

**Conclusion:** Our findings suggest that semaglutide and empagliflozin impact the gut microbial community during treatment. In addition, the baseline gut microbiome can predict semaglutide treatment effects.

## Introduction

Type 2 diabetes (T2D) is a complex disease that is characterized by elevated circulating glucose levels, which can lead to a significant reduction in the quality of life and severe complications such as kidney failure, amputations, and premature cardiovascular mortality^1,2^. In 2021, more than 480 million people were living with type 2 diabetes, and its prevalence is expected to rise to 700 million by 2040^3^. Therefore, T2D significantly burdens healthcare systems worldwide, and effective treatment options are continuously being developed^1^. Despite these novel antidiabetic medications improving T2D treatment results, their efficacy may decrease over time and could be intolerable due to side effects^4–6^. Therefore, understanding the reasons for such inter-individual variability in the treatment response and guiding treatment personalization are necessary.

The gut microbiome can be one of the contributors to the interindividual variability in drug response^7^. The gut microbiome interacts closely with medications, affecting their properties and actions. Such interactions, for example, may involve drug metabolization, which results in altered concentration of the medication and its metabolites or unexpected effects that can potentially affect the overall outcomes or cause severe side effects, leading to treatment discontinuation^8–10^. On the other hand, drug-microbiome interactions are reciprocal. Many drugs, such as antidepressants, proton pump inhibitors, and antipsychotics, have been demonstrated to modulate the gut microbiome^11–13^. These drug-bug interactions have been well characterized for metformin, a common first-choice antidiabetic drug^14^. It has been shown that metformin alters the microbiome composition, but the microbiome is also a part of the mode of action of the drug and can modulate drug efficiency and toxicity^14–15^. Similarly to the first-choice metformin, the action of novel glucagon-like peptide-1 receptor agonists (GLP-1-RA) and sodium-glucose cotransporter-2 inhibitors (SGLT-2i) toward the microbiome has been previously studied^16–20^. However, these studies have shown conflicting results regarding the changes in microbial diversity and abundance of microbial taxa. Therefore, the effect of GLP-1 receptor agonists and SGLT-2 inhibitors on the microbiome and the effect of the microbiome on the drug effect remains to be determined.

Here, we aim to investigate the interactions between the usage of semaglutide (e.g., Ozempic^®^, GLP-1-RA), empagliflozin (e.g., Jardiance^®^, SGLT-2i), and the gut microbiome. Based on the clinical outcomes and gut microbiome measured before and after treatment initiation, we characterize the changes in the microbiome attributable to GLP-1 receptor agonist and SGLT-2 inhibitor admission. In addition, we show that the baseline microbiome can predict changes in the clinical parameters after treatment initiation, suggesting that the microbiome can be used for treatment personalization.

## Methods

### Study design and sample collection

Twenty subjects (10 women and 10 men) were enrolled in the study between November 2019 and January 2023 from the University of Tartu Hospital, Tartu, Estonia. The study followed approval from the Research Ethics Committee of the University of Tartu (290/T-20), and informed consent was obtained from each subject. All participants had been diagnosed with type 2 diabetes, had a BMI of 32 or more, glycohemoglobin (HbA_1c_) <86 mmol/mol (<10%), and were taking 1.5 grams of metformin per day (**Figure 1A**). The main exclusion criteria included the following:

1. having positive glutamic acid decarboxylase (GAD 65) antibodies,
2. an earlier prescription of oral or intravenous antibacterial medications within the last two months,
3. a change in antidiabetic treatment regimen within the last 90 days,
4. previous use of GLP-1 receptor agonists or SGLT-2 inhibitors,
5. use of spironolactone within the last 60 days,
6. current usage of immunomodulating medications (glucocorticoids, cytostatic drugs, or biopharmaceuticals),
7. being pregnant or planning pregnancy,
8. consuming oral contraceptives or oral hormone replacement therapy medications,
9. having previously been diagnosed with a malignant tumor,
10. having a diagnosis of New York Heart Association (NYHA) class III and IV heart failure or a diagnosis of severe liver disease.

**Figure 1.**
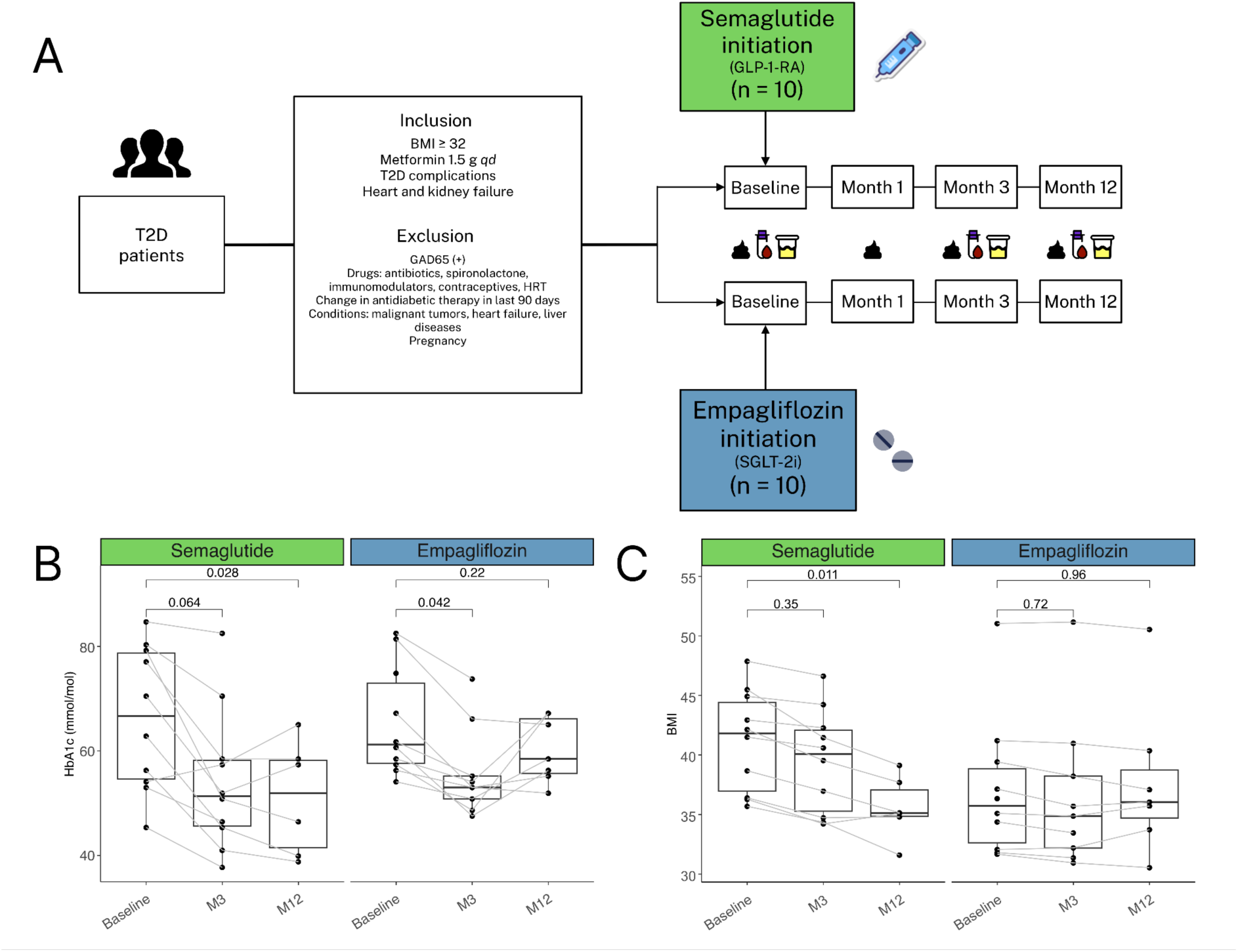
Study design and clinical endpoints. A - overview of the study design. B - Changes in HbA_1c_ and BMI after 3 and 12 months following semaglutide (GLP-1-RA) and empagliflozin (SGLT-2i) initiation. After inclusion and exclusion criteria, 20 T2D patients were recruited to receive either semaglutide or empagliflozin. Subjects provided 4 samples: before treatment initiation and 1, 3, and 12 months after treatment initiation. T2D - type 2 diabetes; HRT - hormone replacement therapy; GAD65 - glutamic acid decarboxylase 65 antibodies, *qd* - daily (*quaque die*).

As a result, the study group taking semaglutide included 6 women and 4 men with ages ranging from 48 to 77 (mean 65.7 ± 8.4) and BMI ranging from 35.69 to 47.87 kg/m^2^ (mean 42.2 ± 5.27). The group prescribed empagliflozin included 4 women and 6 men with age ranging from 52 to 75 (mean 61.7 ± 7.7) and with BMI ranging from 31.67 to 51.04 kg/m^2^ (mean 37.02 ± 5.89) (**Supplementary Table 1**).

The study workflow is shown in **Figure 1**. Each participant donated a venous blood sample, an overnight urine sample, and a fecal sample for microbiome analysis at baseline before treatment initiation, after 1 month, 3 months, and 12 months of treatment. This means that each participant served as their own control, allowing us to minimize inter-individual variability. In addition, body anthropometric measures were recorded. The dose of empagliflozin was 10 mg once a day. A once-weekly subcutaneous (s/c) semaglutide was started at 0.25 mg dose and then up-titrated to 0.5 mg once a week at week 5 and 1 mg once a week at week 9. Treatment efficacy was assessed by measuring glycohemoglobin (HbA_1c_), fasting blood glucose, body mass index (BMI), waist circumference, and inflammation markers (neutrophils, lymphocytes, white blood cells, and neutrophil-to-lymphocyte ratio at Month 3 and Month 12. Several other parameters, like glomerular filtration rate (eGFR), blood lipids, and transaminases, were also analyzed.

### DNA extraction and sequencing

For DNA extraction, study participants collected fecal samples collected with a special stool sampling kit with instructions. Samples were collected using an RNALater sample stabilizing solution and taken to the University of Tartu Hospital, Tartu, Estonia. Immediately after arrival, samples were stored at -20°C and then taken to the Institute of Genomics, University of Tartu, Estonia, where samples were stored at -80°C before follow-up analyses.

DNA extraction of fecal samples was carried out with Qiagen DNeasy PowerSoil^®^ Pro extraction kit (Qiagen, Venlo, Netherlands). Samples were first brought out to room temperature at 25°C for complete dethawing, and 100 µl of starting material was taken from samples. In the following step, 100 µl of Qiagen PowerBead Pro Tube silica gel beads were added to the sample material and heated at 37°C to ensure complete dissolution of salt crystals in RNALater. Next, 800 µl of solution CD1 was transferred to samples following incubation at 65°C for 10 minutes to activate lysis processes of microbial and human intestine cells. The sample disruption step was completed using a tissue homogenizer Precellys^®^ 24 (Bertin Instruments, Montigny-le-Bretonneux, France) at 2500 rpm for 2 x 30 seconds with a 30-second pause between two cycles. The following extraction process was conducted according to the manufacturer’s protocols. All extracted DNA samples were stored at -20°C.

The sequencing process of genomic DNA was conducted twice at the Core Facility of Genomics, Institute of Genomics, University of Tartu, Tartu, Estonia, first in February 2022 (N = 52 samples) and then in January 2023 (N = 26 samples). The concentrations and quality of extracted DNA samples were first assessed using a Qubit^®^ 2.0 Fluorometer (Invitrogen, Grand Island, USA). Extracted genomic DNA was then amplified in the PCR amplification reaction with forward and reverse primers designed for the approximately 460 bp long V3-V4 hypervariable region of the prokaryotic 16S ribosomal RNA gene – 16S_F (5′-TCGTCGGCAGCGTCAGATGTGTATAAGAGACAGCCTACGGGNGGCWGCAG-3′) and 16S_R (5′-GTCTCGTGGGCTCGGAGATGTGTATAAGAGACAGGACTACHVGGGTATCTAAT CC -3)^21^.

### Microbial community analysis

Raw sequences were demultiplexed with Illumina bcl2fastq2 Conversion Software v2.20. Bioinformatic analyses of genomic DNA reads were conducted with the open-source software platform QIIME2 (version 2021.11)^22^. For importing data into QIIME2, the script q2-tools was used in the format of PairedEndFastqManifestPhred33. The first analysis involved denoising sequences with DADA2 software as part of the QIIME2 platform^23^. In addition, forward and reverse sequences were joined at this step. As a result of the first quality control, each base pair position was given a quality score according to whether reads were trimmed and truncated in the following analysis. Due to the better overall quality of reads, the remaining analysis was conducted using forward reads trimmed and truncated at positions 15 and 288. Reads marked as chimeric were removed with the consensus filter. Following the quality control, reads were aligned to amplicon sequence variants (ASV) with software MAFFT^24^. The taxonomic classification was carried out using the QIIME2 script q2-feature-classifier with pre-trained naïve Bayes classifier against the SILVA 16S V3-V4 v132_99 with a similarity threshold set to 99%^25^.

### Statistical analysis

Statistical analyses of microbial ASVs were conducted in R (version 4.3.1). Packages used in the analyses included *mia* (v.1.8.0), *dplyr* (v.1.1.3), *tidyr* (v.1.3.0), and *purrr* (v.1.0.2). Figures were obtained with the package *miaViz* (v.1.8.0), *pheatmap* (v.1.0.12), and *ggplot2* (v.3.4.3). Additional customizations were done using vector graphics software *Inkscape* (v.1.3).

Initially, 78 samples from 21 patients were included in the microbiome analysis, but four samples from one individual were removed due to the missing baseline sample. Additionally, one sample was excluded due to the low number of reads. Thus, the final number of samples was 73 samples from 20 patients.

Alpha diversity was assessed using the *mia* package from microbial genus-level data. The alpha diversity indices included the number of observed genera, the Shannon diversity index, and Pielou’s evenness index. Beta diversity was assessed using five principal components (PC1-PC5) of the principal component analysis on the Aitchison distance. In addition to the diversity analysis, the abundance of genera present in at least 10 samples per study group was analyzed. As a result, we investigated changes in CLR-values of 128 genera in the semaglutide study group and 147 genera in the empagliflozin study group.

To study the effect of two diabetes medications on the gut microbiome, we analyzed the changes in the microbial diversity and the abundance of the prevalent genera after treatment initiation. Changes in the microbiome were assessed with repeated measures ANOVA. Benjamini-Hochberg false discovery rate (FDR) method was used to account for multiple testing^26^. In addition, two-sided dependent Welch t-tests were performed as *post-hoc* tests to study changes in the abundance of genera between two timepoints. *Post-hoc* tests were carried out for genera that showed a difference in the abundance of genera in the repeated measures ANOVA analysis.

To understand whether baseline gut microbiome could predict the efficacy of semaglutide and empagliflozin in antidiabetic treatment, baseline microbiome values were correlated with the change of clinical markers by the third treatment month. This is the timepoint where we detected the largest treatment effect of semaglutide and empagliflozin therapy. The baseline gut microbiome values included alpha and beta diversity indices and CLR-transformed values of the prevalent genera. Pearson correlation coefficient was calculated, and the Benjamini-Hochberg procedure for adjusting the p-values was used to account for multiple testing (FDR < 0.05).

## Results

### Cohort overview and study design

In this study, we recruited 20 subjects with T2D who had been previously using metformin as a part of the treatment regime and had a BMI of at least 32. As the study was conducted in a routine clinical setting, the patients who had more severe obesity and/or diabetes compensation were more likely to be prescribed semaglutide, and subjects with heart and kidney failure or T2D complications were more likely prescribed empagliflozin as a supplementary treatment. Participants were asked to donate microbiome fecal samples four times – at baseline, 1 month (M1), 3 months (M3), and 12 months (M12). Fasting blood samples, overnight urine samples as well as clinical anthropometric measurements were collected at baseline, 3 months (M3) and 12 months (M12) (**Figure 1A**). The microbiome was profiled from fecal samples by sequencing the V3-V4 regions of the 16S rRNA gene. In total, 73 fecal samples were analyzed, with 10 participants taking semaglutide (35 fecal samples) and 10 participants taking empagliflozin (38 fecal samples).

In the semaglutide study group, mean glycohemoglobin (HbA_1c_) values decreased by -11.37 mmol/mol (1.11%) between treatment initiation and the third month (**Figure 1B**, **Supplementary Table 1**). The decrease for empagliflozin users was -13.88 mmol/mol (0.88%). At M12, a decrease in HbA_1c_ constituted -8.2 mmol/mol (1.4%) and -18.25 mmol/mol (0.48%) for semaglutide and empagliflozin users, respectively.

Compared to baseline, BMI (kg/m^2^) for the semaglutide users decreased by 1.69 units by the third treatment month. The BMI in the empagliflozin study group decreased by 0.88 units by M3 (**Figure 1C**, **Supplementary Table 1**). At the 12-month timepoint, the BMI on average decreased by 5.61 units in the semaglutide group and increased by 0.7 units in the empagliflozin group.

### Drug initiation shows no direct effects on the changes in the fecal microbiome

To elucidate the effect of antidiabetic drug usage on the gut microbiome, we compared the baseline gut microbiome with the microbiome composition of M1, M3, and M12 (**Figure 1A**) (**Methods**). First, we analyzed changes in the alpha diversity (observed richness, Shannon index, Pielou index) using repeated measures analysis of variance. No significant changes were detected before and after correction for multiple testing (**Figure 2A, B**; **Supplementary Table 2**). Second, we studied shifts in the overall composition (beta diversity) by analyzing changes in the first 5 principal components of the genus-level CLR-abundance profile. We detected nominally significant changes in PC1 in the semaglutide group and PC5 in the empagliflozin group (repeated measures ANOVA, p <= 0.05). *Post-hoc* tests indicated a nominally significant alteration in PC2 between baseline and M3 for empagliflozin users. However, similarly to alpha diversity, no changes remained statistically significant after applying FDR correction (**Figure 2B**, **Supplementary Table 2**).

**Figure 2.**
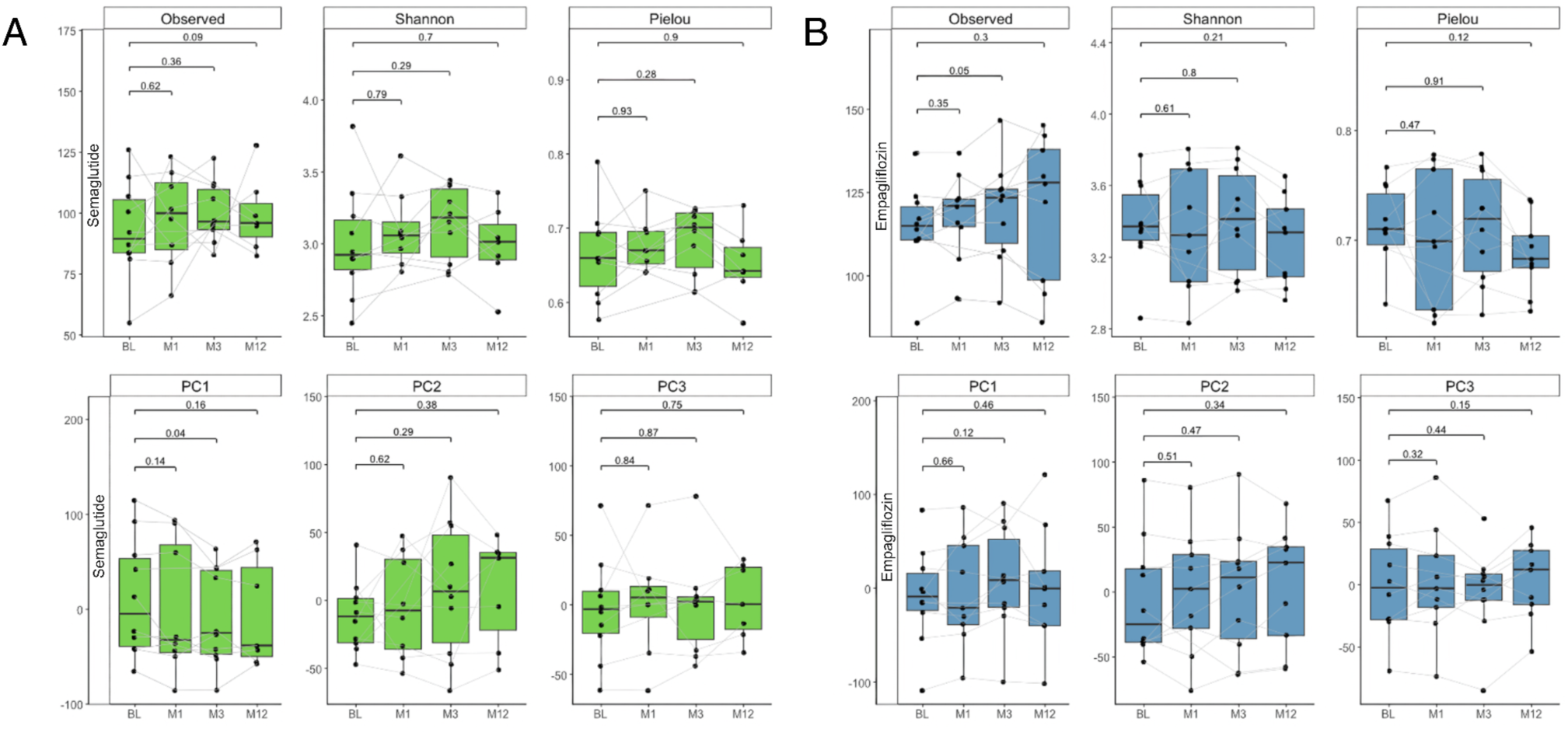
Effect of semaglutide and empagliflozin on the gut microbial diversity in T2D patients. Microbial alpha diversity (observed genera, Shannon diversity, Pielou’s evenness) and three principal components (beta diversity) at four timepoints. Pairwise t-test p-values comparing the microbiome diversity between baseline and later timepoints are shown. A - semaglutide (GLP-1-RA) study group. B - empagliflozin (SGLT-2i) study group. BL - baseline, M1 - 1^st^ month of treatment, M3 - 3^rd^ month, M12 - 12^th^ month.

Next, to characterize the changes in the microbiome in greater detail, we analyzed alterations in the abundance of bacterial genera prevalent in at least 10 samples per study group (128 genera for semaglutide, 147 genera for empagliflozin). First, we aimed to identify genera with altered abundance in between any of the four timepoints. In the semaglutide group, we identified the abundance to be changed between six genera (repeated measures ANOVA, p <= 0.05). These included genera *Actinomyces*, *Candidatus Soleaferrea, Fusobacterium, Haemophilus, Pseudobutyrivibrio,* and *Veillonella* (**Figure 3A**, **Supplementary Table 2**). However, none of these associations remained significant after applying the FDR correction. Similarly, in the empagliflozin group, the abundance of nine bacterial genera was identified to be different between the timepoints (repeated measures ANOVA, p <= 0.05). These genera included Agathobacter, *Akkermansia*, *Coprococcus 2, DTU089*, *Parasutterella*, *Romboutsia, Ruminococcaceae UCG-014*, *Turicibacter*, and uncultured_bacterium 8 from the order Mollicutes RF39 (**Figure 3B, Supplementary Table 2**). However, no significant associations were detected after p-value correction.

**Figure 3.**
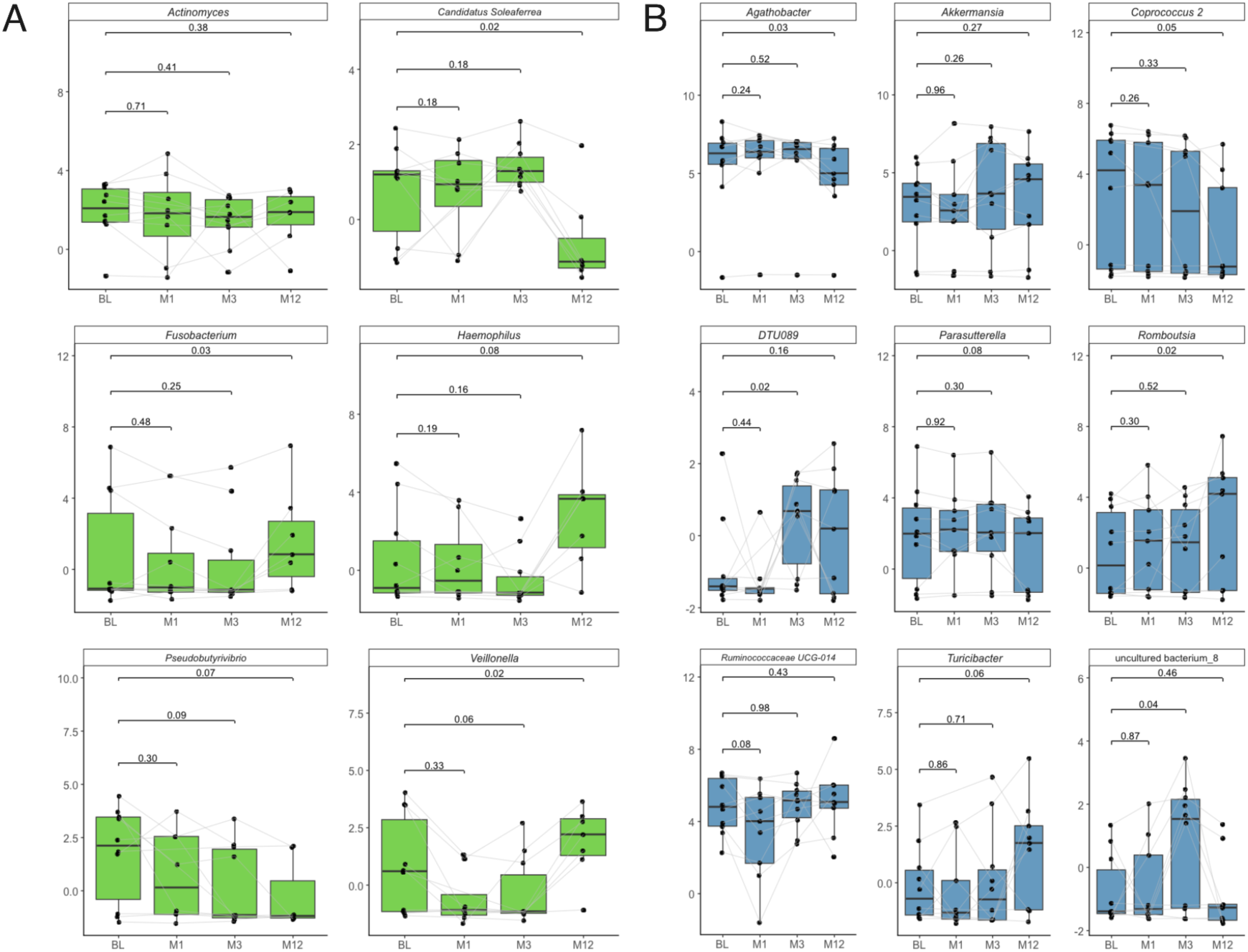
Changes in the CLR-transformed abundances of bacterial genera in the semaglutide and empagliflozin study group. A - semaglutide study group (GLP-1-RA), B - empagliflozin study group (SGLT-2i). Genera detected as nominally significant in the repeated measures ANOVA analysis are shown. The p-values on the plot refer to *post-hoc* analysis using pairwise t-tests to compare the changes between the baseline timepoint and later timepoints. BL - baseline, M1 - 1st month of treatment, M3 - 3rd month, M12 - 12th month.

Next, for the nominal associations identified, we carried out further analyses to identify the timepoints between which the abundance of the genera was changed (*post-hoc* pairwise t-tests). Interestingly, we observed that in both study groups, the changes in the abundances of the bacteria were not evident when comparing the baseline with the latter timepoint. Instead, the change was more evident between the microbiome collected at Month 3 and Month 12 (**Figure 3**, **Supplementary Table 2**). Thus, the results suggest that semaglutide and empagliflozin do not directly affect the gut microbiome. The changes in the microbiome observed between the latter timepoints are likely due to changes in metabolic health or lifestyle.

### Gut microbiome correlates with treatment efficacy

Next, we evaluated the ability of the baseline gut microbiome to predict changes in the major clinical endpoints, indicating the efficacy of the antidiabetic therapy. We calculated the Pearson correlation between the CLR-transformed abundance of the prevalent genera, alpha and beta-diversity metrics, and changes in clinical markers between BL and M3. As for clinical parameters, we focused on BMI, glycohemoglobin (HbA_1c_), measures of lymphocytes and neutrophils, neutrophil-to-lymphocyte ratio (NLR), and amount of white blood cells (WBC). In the empagliflozin study group, glomerular filtration rate (GFR) was additionally included to assess how the baseline microbiome influences the drug’s effect on kidney function. In the semaglutide study group (GLP-1-RA), 55 out of 816 associations were nominally significant (p < 0.05), with the most association with the change in white blood cells and neutrophil-to-lymphocyte ratio (NLR) (**Supplementary Table 2**). After the FDR correction, the correlation between the abundance of *Alistipes* and neutrophil and NLR change remained significant (**Supplementary Table 2**). A higher abundance of *Alistipes* was associated with a higher decrease in neutrophils and NLR. Most importantly, six genera were found to be associated with the change in HbA_1c_ (**Figure 4A**, **Supplementary Table 2**). In the empagliflozin group, 48 associations among 1085 were detected as nominally significant (p < 0.05) (**Figure 4B**, **Supplementary Table 2**). However, none of these correlations were significant after correcting for multiple testing. Similarly to the semaglutide study group, the associations between six bacterial genera and the change of Hb_A1c_ were found to be nominally significant, including genera *Prevotellaceae NK3B31 group*, *Candidatus Soleaferrea*, and *Escherichia-Shigella* (**Figure 4B**, **Supplementary Table 2**). For example, a higher abundance of *Escherichia-Shigella* indicates a higher reduction in HbA_1c_. Therefore, our results suggest that the fecal microbiome could be used to predict the treatment outcome of semaglutide and empagliflozin.

**Figure 4.**
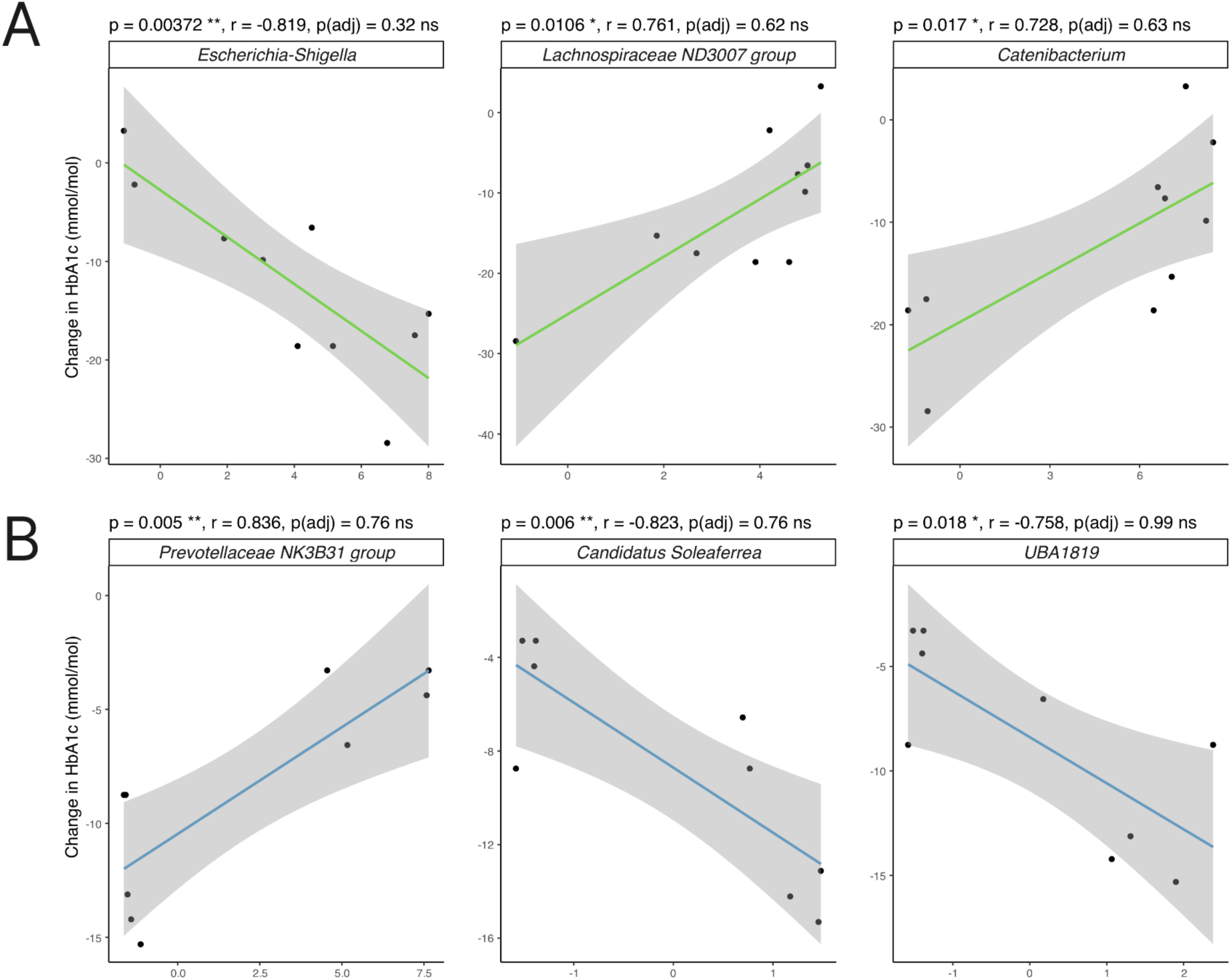
Three nominally most significant associations between the baseline microbiome and change in glycohemoglobin (HbA_1c_) by the third treatment month. A - semaglutide study group (GLP-1-RA), B - empagliflozin study group (SGLT-2i). Associations were estimated using Pearson correlation coefficients.

## Discussion

In this study, we investigated the effect of two novel T2D medications – semaglutide and empagliflozin – on the gut microbiome of T2D patients and evaluated the possibility of using gut microbiome for predicting the efficacy of T2D treatment. To our knowledge, among the GLP-1 receptor agonists, this is the first study to analyze changes in the microbiome after semaglutide initiation. We found that semaglutide and empagliflozin have no significant effect on gut microbial composition. However, we found the baseline microbiome to correlate with the change in clinical outcomes. The associations that emerged from the correlation analysis may pinpoint the gut microbiome’s potential capacity to aid in personalizing antidiabetic therapy.

Surprisingly, we found no significant effect of semaglutide and empagliflozin on the gut microbial alpha and beta diversity. These findings agree with several previous studies, where gut microbiome diversity remained unaltered following GLP-1 receptor agonist or SGLT-2 inhibitor consumption throughout the treatment period^17,16^. However, Deng *et al.* detected an increase in gut microbial alpha diversity when investigating the effect of empagliflozin^19^. Nevertheless, they compared the alpha diversity between naive empagliflozin initiators with the alpha diversity of metformin users. In contrast, subjects in our study had already been taking metformin before the treatment with empagliflozin. The actions of metformin on the gut microbial community have been previously well-characterized and, thus, could potentially obscure the potential effects of additional semaglutide and empagliflozin consumption^14^.

Previous studies have investigated the associations between the gut microbiome and antidiabetic treatment efficacy. For example, Tsai *et al.* detected 17 distinct microbial features associated with alterations in HbA_1c_ for subjects taking liraglutide or dulaglutide, two glucagon-like peptide-1 receptor agonists^17^. Interestingly, some of the bacterial features overlapped with the results of our study, namely the genera *Bacteroides* and *Lachnoclostridium*. For sodium-glucose cotransporter-2 inhibitors, earlier studies have primarily analyzed the associations between the gut microbiome and the efficacy of dapagliflozin or gliclazide^18^. Unlike the studies that suggest the effect of microbiome on the efficacy of glucagon-like peptide-1 receptor agonists, the ability of baseline gut microbiome to predict the response to sodium-glucose cotransporter-2 inhibitors remains to be demonstrated. Nevertheless, we are unaware of any studies that have shown the effect of microbiome on the efficacy of semaglutide. Based on previous results and findings from our study, the potential links between baseline gut microbiome and glucagon-like peptide-1 receptor agonists merit further investigation in personalizing T2D treatment.

Considering that most medications affect the gut microbiome over a more extended period^27^, the design of this study allows us to define more clearly the changes in microbial composition and diversity. Compared to previous studies on T2D medications and gut microbiome that have implemented only a single or two timepoint sample and data collection, subjects in our study were monitored at four timepoints for 12 months. Taking advantage of the long follow-up period allowed us to show that the changes in the microbiome composition are not observable directly after treatment initiation but after a more extended time. Regarding this, future studies that monitor longitudinal alterations in the gut microbial community should be able to detect precise changes in the microbiome and more accurately predict the effect of antidiabetic drug action using gut microbial data.

It should be noted that several technical aspects could affect the statistical power of analysis and interpretation of the results, e.g., relatively small cohort size and missing samples at various timepoints for some participants. In addition, it is worth reminding that while the consumption of semaglutide or empagliflozin may have caused some direct changes in the abundance of these bacterial genera, the general effect of T2D treatment might have indirectly altered the compositional values of bacterial taxa, e.g., through reduced hyperglycemia and body mass index.

In summary, the findings from our study show that the gut microbial community can predict semaglutide treatment effects based on changes in clinical markers. In addition, semaglutide and empagliflozin are associated with gut microbiome alterations throughout antidiabetic therapy, even if changes arose due to the indirect effects of T2D treatment, e.g., decreased BMI and HbA_1c_. The ability of the microbiome to predict treatment efficacy warrants further investigation into the causality and functional significance of these findings. This could result in prescribing the most suitable T2D medication before treatment or altering the gut microbial community during treatment. Overall, the longitudinal data of our study provide the means to undertake further studies to validate these signals in larger cohorts.

## Supporting information

Supplementary Table 2. Analysis of bidirectional effects between T2D medications and gut microbiome

Supplementary Table 1. Overview of the study cohort

## Data availability

The data generated in this study have been deposited in the European Genome-Phenome Archive database (https://www.ebi.ac.uk/ega/) under accession code xxx. This study was approved by the Research Ethics Committee of the University of Tartu (approval No. 290/T-20).

## Acknowledgments

This study was funded by the European Union through the European Regional Development Fund Project No. 2014-2020.4.01.15-0012 GENTRANSMED. Data analysis was carried out in part in the High-Performance Computing Center of the University of Tartu. We thank Reidar Andreson for the bioinformatic support.

## Author Contributions

O.A., E.O., and V.V. conceptualized and supervised the study. I.R. and V.V. recruited study participants and organized sample collection and clinical monitoring. K.L.K. and A.K. performed DNA extraction and prepared metadata for microbiome analysis. A.K. performed the data analysis. A.K. and O.A. interpreted the data, prepared the figures, and wrote the manuscript. All authors read and approved the final paper.

